# Single-Shot Lightweight Model For The Detection of Lesions And The Prediction of COVID-19 From Chest CT Scans

**DOI:** 10.1101/2020.12.01.20241786

**Authors:** Aram Ter-Sarkisov

## Abstract

We introduce a lightweight model based on Mask R-CNN with ResNet18 and ResNet34 backbone models that segments lesions and predicts COVID-19 from chest CT scans in a single shot. The model requires a small dataset to train: 650 images for the segmentation branch and 3000 for the classification branch, and it is evaluated on 21292 images to achieve a **42**.**45%** average precision (main MS COCO criterion) on the segmentation test split (100 images), **93**.**00%** COVID-19 sensitivity and F1-score of **96**.**76%** on the classification test split (21192 images) across 3 classes: COVID-19, Common Pneumonia and Control/Negative. The full source code, models and pretrained weights are available on https://github.com/AlexTS1980/COVID-Single-Shot-Model.

## 1 Introduction

Examination of chest CT scans is one of the most popular and accurate ways of predicting COVID-19, alongside x-ray radiography and real-time polymerise chain reaction (RT-PCR): it is faster than RT-PCR and more accurate than x-ray. Depending on the stage of the virus, it can have higher sensitivity to COVID-19 than RT-PCR ([PCP20, AYH^+^20]. Since the onset of the COVID-19 a large number of convolutional neural networks (convnets) and other deep learning (DL) models for the detection of COVID-19 from chest CT scans and segmentation of lesions were introduced. The classifiers typically use a feature extractor like ResNet with a problem-specific logit output (e.g. COVID-19 vs Common Pneumonia), like in [GWW20, LQX^+^20, PCP20, BGCB20]. Most segmentation models use a semantic segmentation model like U-Net to predict the classes’ masks at a pixel level, e.g. [FZJ^+^20, ZLZ^+^20], or to augment the classifier by concatenating the mask prediction with deep features, e.g. in [WGM^+^20].

Recently introduced model COVID-CT-Mask-Net [TS20a] augments Mask R-CNN and Faster R-CNN [HGDG17, RHGS15] object detection/segmentation models with a classification module that exploits the detectors’ capacity for predicting bounding boxes and masks of the regions of interest (RoIs) equipped with normalized (confidence) scores, expressing the model’s ‘opinion’ about the class of the object. The batch of these predictions is converted into a vector of features that the classification module in COVID-CT-Mask-Net must learn. In [TS20b, TS20c] a number of modifications were presented, including truncated lightweight versions of the base model that achieve a similar accuracy of COVID-19 prediction and lesion segmentation. The main drawback of this approach is that it consists of two stages: first, a segmentation model (Mask R-CNN) is trained on the data with ground truth (gt) masks, then, a classification model is derived from it and trained on the data labeled at the image level (COVID-19, Common Pneumonia, Control).

In this paper we present a solution that trains a model to segment lesions in chest CT scans and predict the class of the image (COVID-19, Common Pneumonia, Control) in a single shot (single-shot model, SSM). The solution relies on Mask R-CNN’s capacity for detecting objects and segmenting their masks. Mask R-CNN consists of four main stages: backbone (ResNet)+Feature Pyramid Network (FPN), Region Proposal Network (RPN), Region of Interest (RoI) and mask prediction. We evaluate two approaches: with Mask R-CNN pretraining and without any pretraining. For the classification part, we explore three approaches: first, we reuse the segmentation branch of RoI to create a batch for the image classification, which is in line with the solution in [TS20a], second, we augment RoI module with a parallel classification branch that outputs RoIs for the image classifier; finally, we abandon the pretraining stage altogether, and adapt both branches simultaneously from scratch.

The single-shot model has a number of advantages:

- This approach solves both lesion detection/segmentation and COVID-19 vs Common Pneumonia vs Control class prediction from chest CT scans in a single shot, as pretraining is not necessary.
- The models are lightweight (less than 10*M* parameters), as a result, training and evaluation are very fast. On a CPU, processing of a single 512*×*512 CT scan slice takes between 3.81-7.08s, which includes the full segmentation output and the image class prediction.
- The model can be easily adapted to new data.

To the best of our knowledge, this is the first paper that presents a single-shot solution for both lesion instance segmentation and COVID-19 classification. The rest of the paper is structured as following: in Section 2 we discuss the datasets for both problems, in Section 3 we present the methodology, in Section 4 we discuss the experimental setup and the results, Section 5 concludes.

## 2 Data

We require two separate sets of the training data: segmentation data and classification data. Both of these sets are taken from CNCB-NCOV [ZLS^+^20], http://ncov-ai.big.ac.cn/download resource. The segmentation data (750 images labelled at pixel level) is split randomly into 650 training and validation and 100 test images. Masks for the lesion classes, Ground Glass Opactity (GGO) and Consolidation (C) are merged into a single positive lesion class. Clean lung masks are merged with the background. All 750 images (scan slices) were taken from COVID-19-positive patients, but some of the slices are negative (no lesions present). These are skipped during training, and labelled as a single negative observation at test stage.

For the 3-class (COVID19, CP, Normal) classification data we use the COVIDx-CT train/test/validation splits, [GWW20]. We use the same sample of 3000 images (1000/class) from the train split as in [TS20c], and also used the test and validation splits in full, see Table 1. The splits are consistent across classes and patients. This means that negative slices taken from the positive (COVID-19 or CP) patients were removed from the data altogether, and only those with lesions were kept, and every patient was randomly assigned into only one of the splits, [GWW20]. This is one of the key advantage of SSM: it generalizes very well to the unseen data while using only a small portion of the training dataset.

**Table 1:**
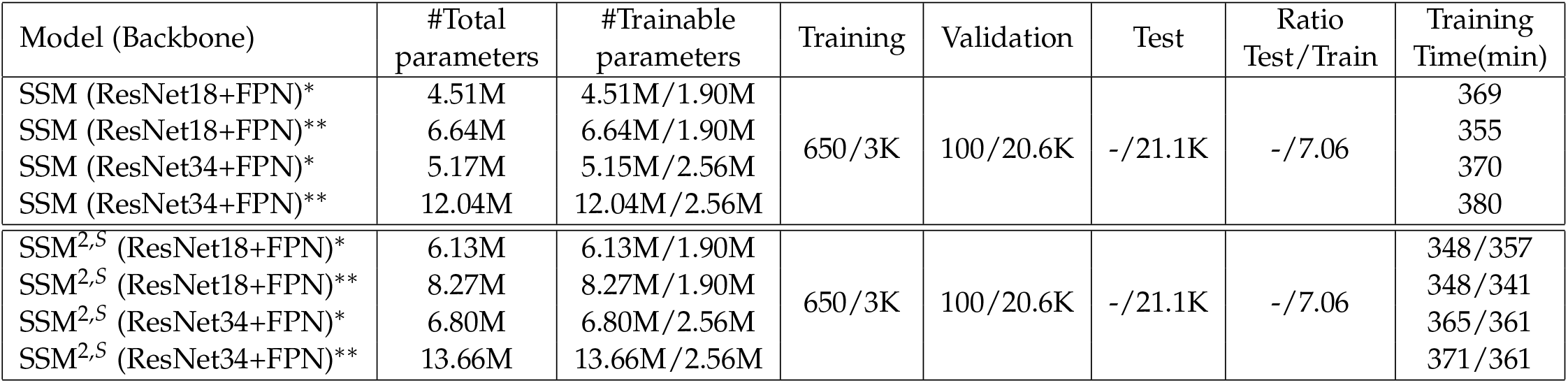
Comparison of models’ sizes and training datasets. The #trainable parameters is for the segmentation/classification problem. Models with ^∗^ have two backbone blocks, models with ^∗∗^ have three backbone blocks. Models with ^2^ superscript have two parallel feature blocks in the RoI layer, models with the ^*S*^ superscript were trained from scratch. Models trained from scratch have the same number of parameters as SSM^2^. Training time for SSM^2,*S*^ are pretrained/trained from scratch. The training time for all pretrained models includes the pretraining.

| Model (Backbone) | #Total parameters | #Trainable parameters | Training | Validation | Test | Ratio Test/Train | Training Time(min) |
| --- | --- | --- | --- | --- | --- | --- | --- |
| SSM (ResNet18+FPN)* | 4.51M | 4.51M/1.90M | 650/3K | 100/20.6K | -/21.1K | -/7.06 | 369 |
| SSM (ResNet18+FPN)** | 6.64M | 6.64M/1.90M |  |  |  |  | 355 |
| SSM (ResNet34+FPN)* | 5.17M | 5.15M/2.56M |  |  |  |  | 370 |
| SSM (ResNet34+FPN)** | 12.04M | 12.04M/2.56M |  |  |  |  | 380 |
| SSM<sup>2,S</sup> (ResNet18+FPN)* | 6.13M | 6.13M/1.90M | 650/3K | 100/20.6K | -/21.1K | -/7.06 | 348/357 |
| SSM<sup>2,S</sup> (ResNet18+FPN)** | 8.27M | 8.27M/1.90M |  |  |  |  | 348/341 |
| SSM<sup>2,S</sup> (ResNet34+FPN)* | 6.80M | 6.80M/2.56M |  |  |  |  | 365/361 |
| SSM<sup>2,S</sup> (ResNet34+FPN)** | 13.66M | 13.66M/2.56M |  |  |  |  | 371/361 |

## 3 COVID-19 Single-Shot Segmentation And Classification Model

Mask R-CNN is one of the state-of-the-art models that detects and segments separate objects in images using Region Proposal Net (RPN) and Region of Interest Net (RoI), which is different to semantic segmentation predicting classes at a pixel level, such as FCN [LSD15] and UNet [RFB15]. Unlike semantic segmentation models, Mask R-CNN ‘understands’ separate objects, together with their labels and masks, and is therefore good at handling problems like partial occlusion. Nevertheless, Mask R-CNN does not make *global* predictions, i.e. prediction at an image level (class of the image). Some previous result, e.g. [TS20a, TS20b] extend Mask R-CNN to do global prediction, but in a separate model.

Single-shot model (SSM) that we present in this paper solves both the problem of detecting and segmenting lesions in chest CT scans and predicting whether the scan slice is COVID-19-positive, Common Pneumonia-positive or Negative (no type of Pneumonia). Figure 1 summarizes the SSM training and evaluation mechanism. We present three main approaches to the model’s training:

**Figure 1:**
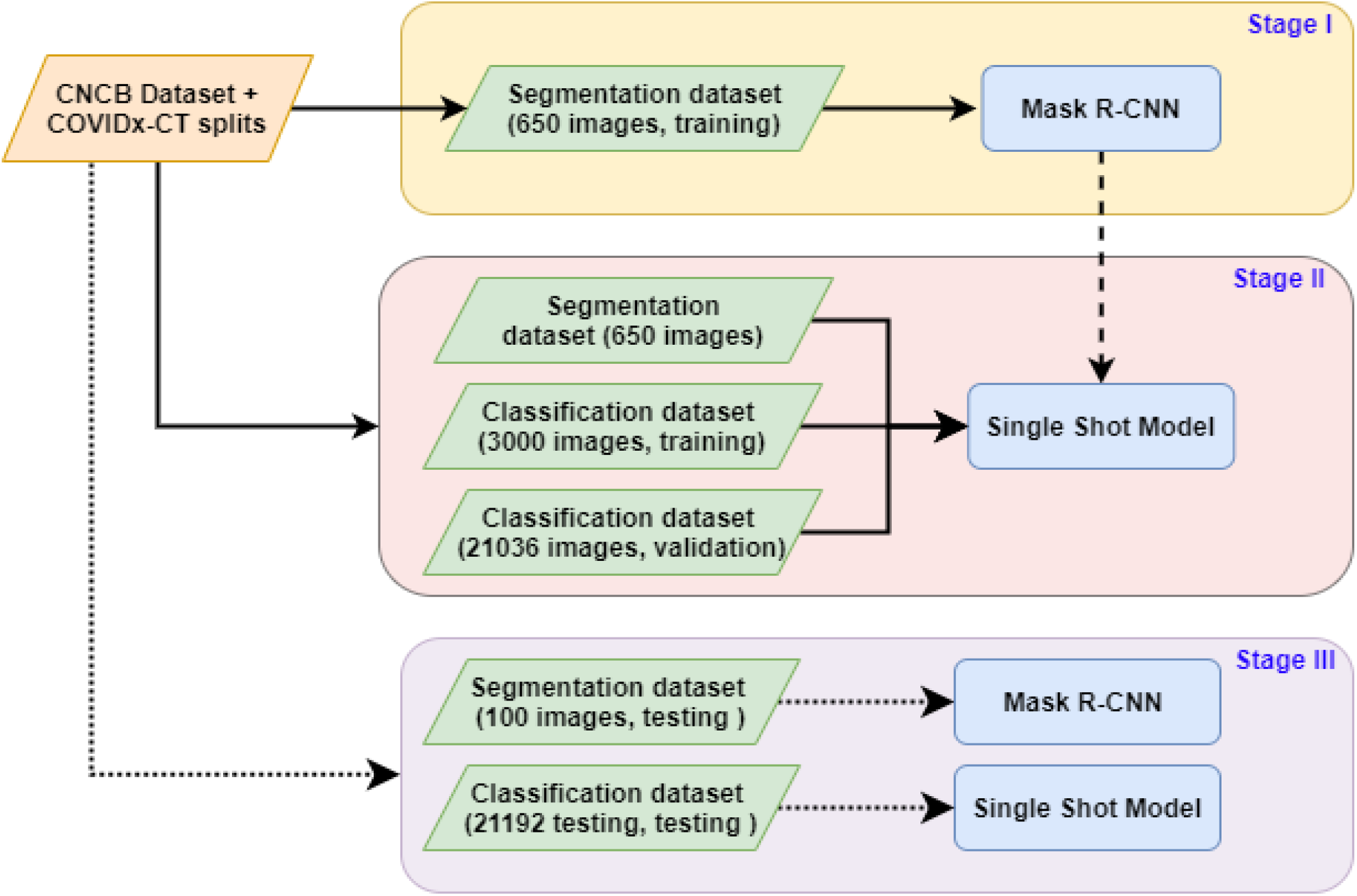
Overview of the method. Black arrows: training/validation data copy or input, dotted arrows: test data, broken arrow: model augmentation and weights copy. Green blocks: data splits, blue blocks: models. Stage I: Segmentation pre-training, Stage II: joint segmentation and classification training, Stage III: model testing. For the The SSM trained from scratch, Stage I is skipped. The order of the operations is top-down.

**Figure 2:**
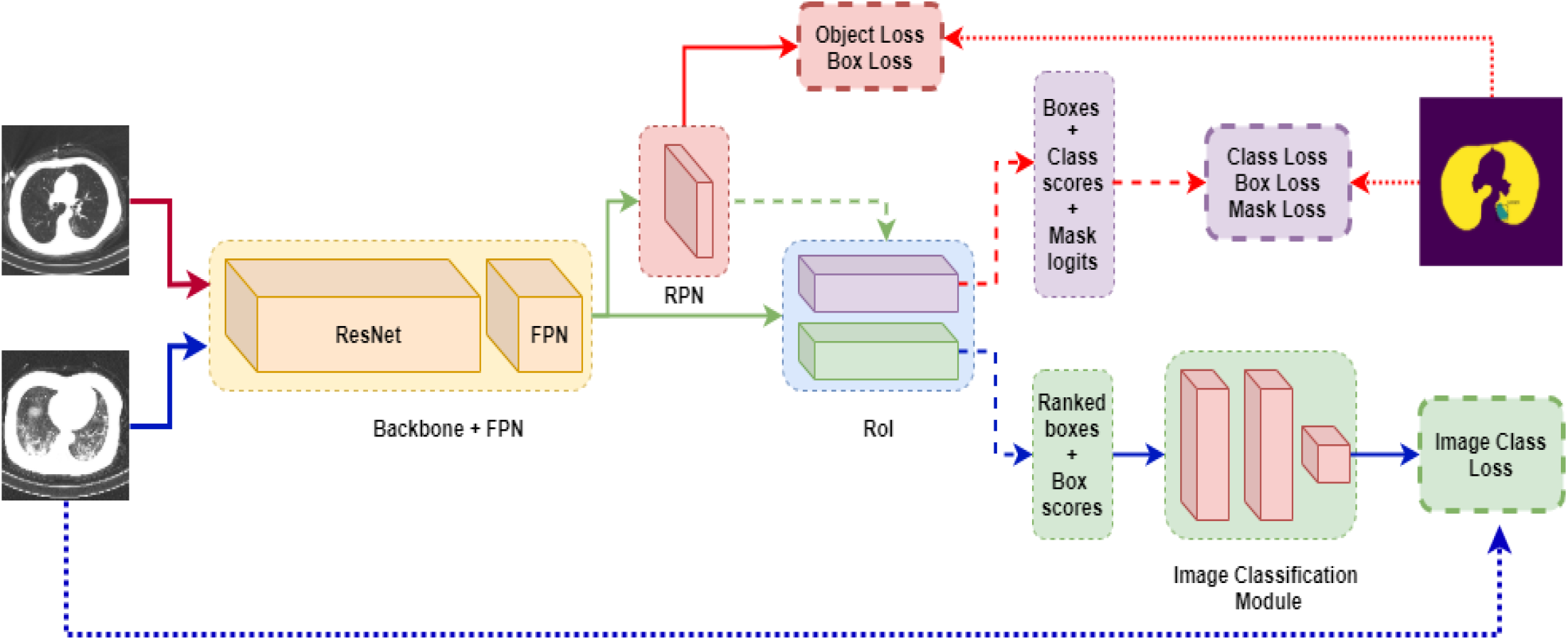
Architecture of the Single Shot Model with two parallel RoI branches. Both segmentation and image classification losses are computed. Green arrows: segmentation and classification stages, red arrows: only segmentation stage, blue arrows: only classification stage. Normal arrows: data/tensors, broken arrows: batches/samples, dotted arrows: labels. Best viewed in color.

1. pretrained segmentation branch + single branch for segmentation+classification, Section 3.2,
2. pretrained segmentation branch + two parallel branches for segmentation+classification, Section 3.2,
3. training from scratch+two parallel branches for segmenation+classification, Section 3.3.

An important idea we use is the encoding of bounding boxes. Mask/Faster R-CNN use several encodings for the bounding box coordinates: (a) encoded ground truth box coordinates for the RPN/RoI loss computation, (b) decoded box offsets for the RPN detections output, (c) decoded RoI detection output for the inference, (d) scaled boxes in (c) to match the image size. For the image classification problem we use the boxes before decoding in (c), i.e. the outputs of the detection branch in RoI. Since the branch was trained using encoded ground truth targets in (a), it outputs box coordinates encoded (normalized) using non-trainable coordinate weights (see Torchvision Mask R-CNN implementation, https://pytorch.org/docs/stable/torchvision).

### 3.1 SSM Loss Function

RPN solves an object vs background binary problem, RoI solves a multiclass problem (objects vs background). Equation 1, loss of the segmentation branch, is the same as in [HGDG17, RHGS15]. Here 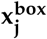 is the bounding box prediction, 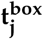 are gt (target) box coordinates, *f*_*j*_(*x*) are the logit scores, *C* is the index of the class of the *j*^*th*^ prediction, e.g. *C* = 0 is the background class. *L*_1_smooth is a variation of absolute distance function, [RHGS15]. Per-class mask labels 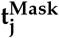 are resized binary masks and predictions for this class 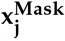 are logit score maps of the same size, so the loss is taken for each mask pixelwise. Therefore RPN object loss and *L*_*Mask*_ are binary cross-entropy (log-sigmoid) loss functions (object vs background), and RoI class loss is a cross-entropy loss functions (log-softmax). The encoding of the ground truth box coordinates is also the same as in [RHGS15]. For the masks loss, only positive predictions in the sample are used 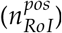, as well as for the box coordinates. For the object and class scores the whole sample (*n*_*RPN*_, *n*_*RoI*_) is used.

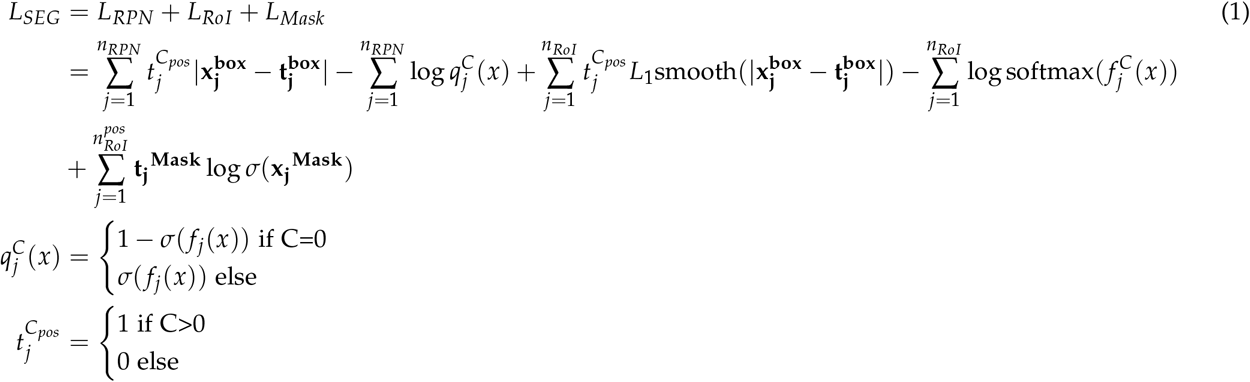

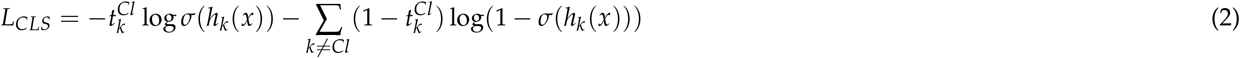

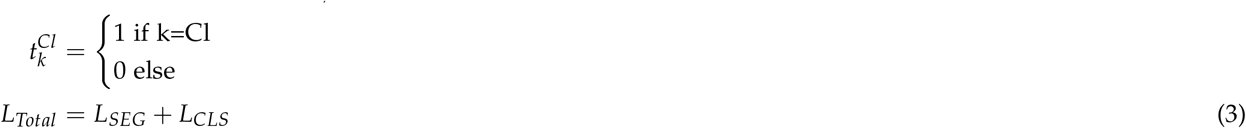

Equation 2 is the only loss function for the classification branch. We use class logits *h*_*k*_(*x*)(*x* is the output from the final logit/score layer in the model), and binary cross-entropy loss functions, *C* is the total number of classes, *Cl* is the index of the correct class. For example, if COVID-19 is the correct class of the image, the label vector is [0, 0, 1], *Cl* = 2. The total loss, Equation 3 is taken without any coefficients adjusting either *L*_*SEG*_ or *L*_*CLS*_.

### 3.2 Weight Sharing With The Pretrained Segmentation Model

The first approach to the SSM is summarized in Figure 1. Mask R-CNN is pretrained on the segmentation data. Then, its weights are copied into the SSM, which then solves both segmentation and classification problems. This pretraining is necessary because the RoI batch output, i.e., the batch input in the image classification module, Figure 3b, is expected to have several important properties (see [TS20a] for details):

**Figure 3:**
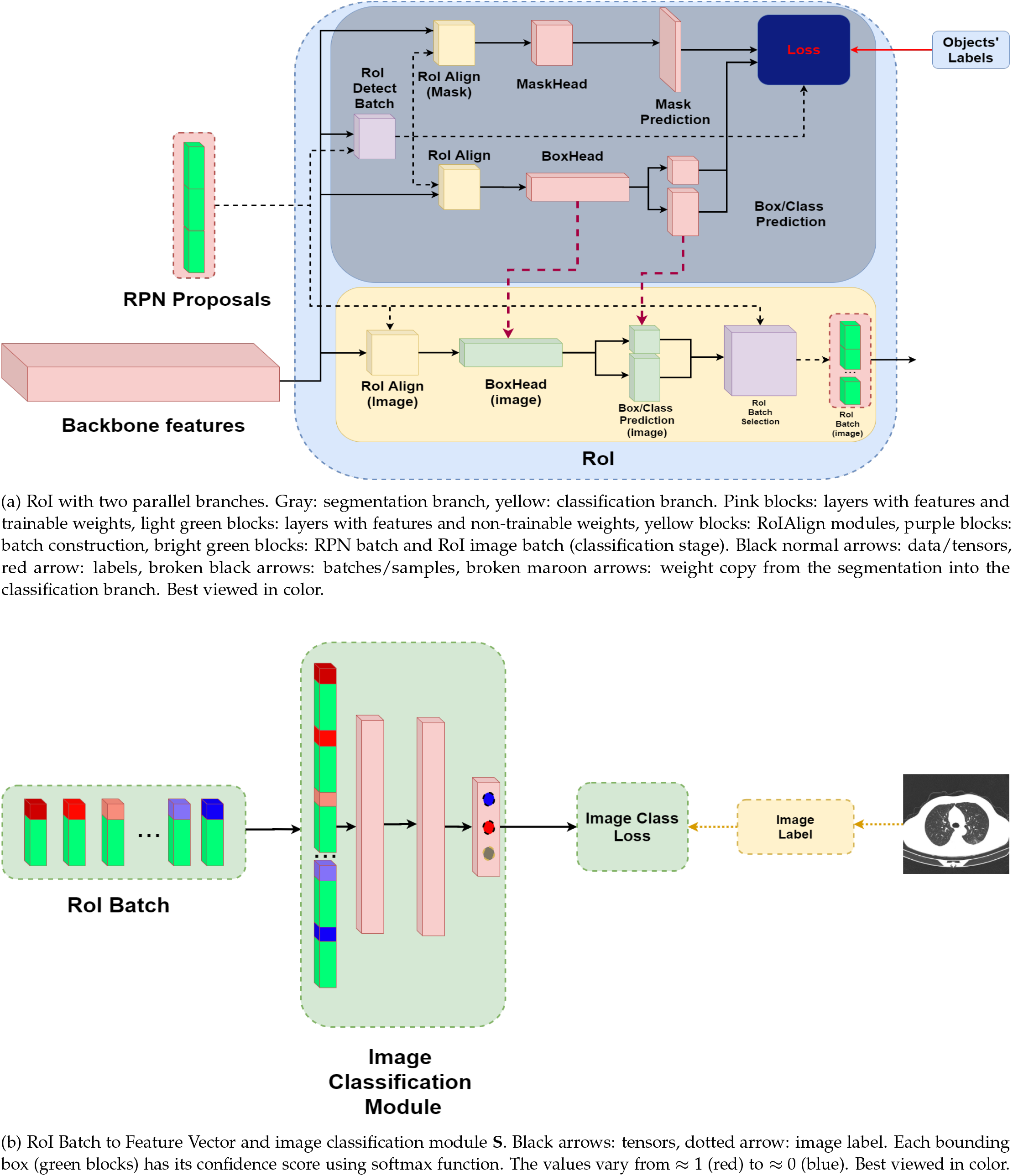
RoI layer with two parallel branches and batch to feature conversion.

1. Contain encoded box coordinates (x, y, height, width),
2. Contain a normalized (softmax filter) confidence score,
3. Elements are ranked in the decreasing order of their confidence scores.

Module **S** is expected to learn this distribution, which applies certain restrictions on the RoI weights for the classification problem: they can only be trained using object-level data (box coordinates, class, mask), which we obviously do not have for the image classification dataset. We therefore resolve this problem in three different ways, each with a different approach to the adaptation of the classification branch.

- Use pretrained RoI weights and the same branch for both problems,
- Use pretrained RoI weights and two parallel branches for each problem,
- Train both branches from scratch, no pretraining.

The last approach is explained in Section 3.3, and the last two approaches are illustrated in Figure 3a. In all approaches, one image of segmentation data follows one image of classification data. The main difference is the classification weights update rule.

In the first approach, the RoI branch weights are shared for both problems, i.e. the only architectural difference between Mask R-CNN and SSM is the **S** module in Figure 3a that is connected to a single RoI branch. At the segmentation stage, all weights (i.e., backbone, RPN, RoI, Masks) except **S** are updated. In the classification stage these weights are frozen, and only **S** weights are updated. That is, RoI branch weights are only updated using segmentation loss. The main difference between segmentation and classification stages is the RoIDetectBatch in Figure 3a that samples the training batch for RoI from RPN output using gt data during the segmentation stage, and accepts the full RPN output during the classification stage, see [TS20a] for the details.

In the second approach, we create a parallel branch in the RoI layer, see Figure 3a with the architecture identical to the existing one (RoIAlign followed by the regional features with two fully connected layers followed by class and box branches). This branch takes the full input from the RPN and outputs the RoI batch for **S**. The most important step here is the selection of the sample of fixed size (RoIBatchSelection layer in Figure 3a), which is described in detail in [TS20a] and is identical to the first approach. The weights in this classification branch are copied from segmentation branch before the start of the algorithm and are never updated.

### 3.3 On-line Weight Sharing With the Segmentation Model

This approach does not require any pretraining at all (incl. backbone weights) and is therefore the easiest to train. Its architecture is identical to the model with two branches in Section 3.2. The main difference is that, since the model is trained from scratch, instead of freezing the weights, we copy the weights from the segmentation branch if there’s an improvement in the segmentation loss in this iteration. In this setup the classification branch outputs the required distribution of boxes without updating its weights wrt the image loss, but instead adapting it alongside the segmentation branch. The weights are copied across all trainable RoI layers: RoI head and the detection branch (class + box coordinates). In this setup RPN also adapts only to the segmentation data, thus maintaining its strength of predicting the objects, which is important for the RoI segmentation branch.

Sampling rules remain the same. First, RPN batch is used in full (without RoIDetectBatch sampling) to create the aligned regional features maps. These are fed through the classification module to output the batch of encoded box coordinates+confidence scores. Then, RoIBatchSelection creates the RoI batch output by removing the overlapping boxes and keeping the batch size fixed and maintaining the balance of high- and low-ranking boxes (see Figure 3b). Thus the RoI batch maintains the characteristics detailed in Section 3.2 essential to the image classification problem and is not affected by the image class loss. Finally, the architecture of **S** is identical across all three approaches: RoI batch is converted to a feature vector (input), followed by two fully connected layers equipped with ReLU activation functions and the final class logits layer.

## 4 Experimental results

We test empirically the following three hypotheses in order to determine the best overall model:

1. Reducing the model’s depth from full (4 layers) to 3 and 2, while keeping a single FPN module (see [TS20c] on the matter of model truncation for this problem),
2. Changing the model’s architecture from ResNet50 to ResNet34 and ResNet18,
3. Comparing three frameworks introduced in Section 3: single branch, separate classification branch with the pretrained frozen weights and two parallel branches trained from scratch.

For the experimental setup we selected two backbones: ResNet18 and ResNet34 becasue in [TS20c] it was shown that smaller models achieve the classification accuracy close to that of the larger models like ResNet50 with just a fraction of the model’s size. It was also shown that truncating models, i.e. removing either the last or the last two blocks (see [HZRS16] for the explanation of the residual architecture and Torchvision model zoo, https://pytorch.org/docs/stable/torchvision/models.html for the implementation we used) in fact improves the predictive quality of the model. As a result, in this setup we skip both ResNet50 and full ResNet18/34 models in favor of smaller truncated versions thereof. We also considered three update rules for the image classification step:

1. Module **S** only,
2. Module **S**+full backbone,
3. Module **S**+batch normalization layers in the backbone.

Approaches 1) and 3) require the least number of weights updates, and 3) is in line with [TS20a]. Nevertheless, with 2) the best results were achieved across all models, hence the results reported in Tables 2 and 3 were attained using rule 2. In total, we trained 12 different variants of the model: 4 with a single (segmentation+classification) RoI branch, 4 with two parallel RoI branches, and 4 with two parallel RoI branches from scratch. In each setup, ResNet18/34+FPN with either 2 or 3 blocks was used as a backbone. Each of the first 8 models was first pretrained with a purely segmentation architecture (see Figure 1). The segmentation model was pretrained for 50 epochs with Adam optimizer, learning rate of 1*e−*5 and weight decay coefficient of 1*e−*3. Important Mask R-CNN hyperparameters such as non-maximum threshold were the same as in [TS20c].

**Table 2:**
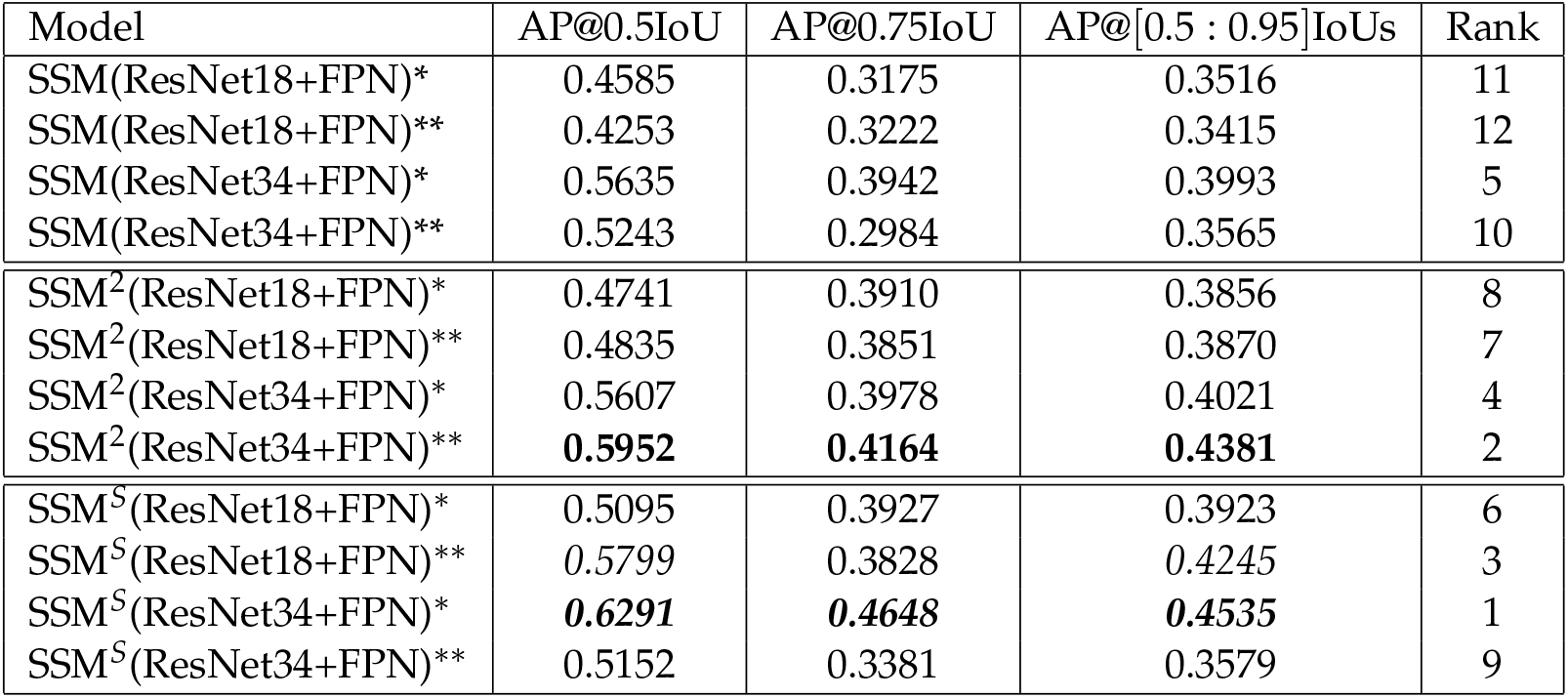
Average precision of segmentation models at three different IoU thresholds. Bold+italicized: best, bold:second-best, italicized: third-best. The rank is based on the AP across all IoU thresholds, which is the main MS COCO 2017 metric.

| Model | AP@0.5IoU | AP@0.75IoU | AP@[0.5 : 0.95]IoUs | Rank |
| --- | --- | --- | --- | --- |
| SSM(ResNet18+FPN)* | 0.4585 | 0.3175 | 0.3516 | 11 |
| SSM(ResNet18+FPN)** | 0.4253 | 0.3222 | 0.3415 | 12 |
| SSM(ResNet34+FPN)* | 0.5635 | 0.3942 | 0.3993 | 5 |
| SSM(ResNet34+FPN)** | 0.5243 | 0.2984 | 0.3565 | 10 |
| SSM<sup>2</sup>(ResNet18+FPN)* | 0.4741 | 0.3910 | 0.3856 | 8 |
| SSM<sup>2</sup>(ResNet18+FPN)** | 0.4835 | 0.3851 | 0.3870 | 7 |
| SSM<sup>2</sup>(ResNet34+FPN)* | 0.5607 | 0.3978 | 0.4021 | 4 |
| SSM<sup>2</sup>(ResNet34+FPN)** | **0.5952** | **0.4164** | **0.4381** | 2 |
| SSM<sup>S</sup>(ResNet18+FPN)* | 0.5095 | 0.3927 | 0.3923 | 6 |
| SSM<sup>S</sup>(ResNet18+FPN)** | 0.5799 | 0.3828 | 0.4245 | 3 |
| SSM<sup>S</sup>(ResNet34+FPN)* | **0.6291** | **0.4648** | **0.4535** | 1 |
| SSM<sup>S</sup>(ResNet34+FPN)** | 0.5152 | 0.3381 | 0.3579 | 9 |

**Table 3:** Class sensitivity, overall accuracy and F1-score results on COVIDx-CT test data for 3 classes (21192 images). Models with ^*∗*^ have two blocks, models with ^*∗∗*^ have three blocks. Models with ^2^ superscript have two parallel feature blocks in the RoI layer, with ^*S*^ superscript were trained from scratch. The rank is based on F1 score.

| Model | COVID-19 | CP | Normal | Overall | F1 score | Rank |
| --- | --- | --- | --- | --- | --- | --- |
| SSM(ResNet18+FPN*) | 90.20% | 89.52% | 89.34% | 89.58% | 0.8969 | 7 |
| SSM(ResNet18+FPN**) | 83.00% | 89.55% | 98.62% | 92.25% | 0.9220 | 5 |
| SSM(ResNet34+FPN*) | 88.70% | 83.35% | 94.54% | 89.43% | 0.8941 | 8 |
| SSM(ResNet34+FPN**) | 87.13% | 96.75% | 88.04% | 91.23% | 0.9138 | 6 |
| SSM<sup>2</sup>(ResNet18+FPN*) | 83.76% | 78.65% | 87.58% | 83.67% | 0.8370 | 11 |
| SSM<sup>2</sup>(ResNet18+FPN**) | 84.38% | 78.22% | 88.76% | 84.18% | 0.8417 | 10 |
| SSM<sup>2</sup>(ResNet34+FPN*) | 85.48% | 81.80% | 79.39% | 81.47% | 0.8166 | 12 |
| SSM<sup>2</sup>(ResNet34+FPN**) | 85.07% | 78.07% | 91.04% | 85.28% | 0.8528 | 9 |
| SSM<sup>S</sup>(ResNet18+FPN*) | **93.16%** | 95.68% | 96.18% | **95.38%** | **0.9542** | 2 |
| SSM<sup>S</sup>(ResNet18+FPN**) | **93.00%** | 96.53% | 98.64% | **96.75%** | **0.9676** | 1 |
| SSM<sup>S</sup>(ResNet34+FPN*) | 89.62% | 89.99% | 96.76% | 92.93% | 0.9293 | 4 |
| SSM<sup>S</sup>(ResNet34+FPN**) | 91.44% | 95.33% | 92.48% | 93.26% | 0.9333 | 3 |

All single shot models were trained in a similar way, with an Adam optimizer, learning rate of 1*e−* 5 and weight decay coefficient of 1*e−*3. For each image in the classification data, an image from the segmentation data was sampled randomly, and the full model (except the RoI classification branch + image classification module **S**) was updated. Then, the image from the classification dataset was fedforward. Each model was trained for 50 epochs (classification dataset).

Although there’s no metric that balances detection and classification results, from the ones presented in Table 2 and 3 we can infer that the models trained from scratch with the adapted classification branch perform best in classification, including COVID-19 sensitivity and F1 score, and they also perform best and second-best for classification, measured by the mean AP. For the classification problem, adding the third block gives a stable improvement in F1 score for each variant, but this effect is not clear for the segmentation problem. At the same time, switching from ResNet18 to ResNet34 with the same number of blocks tends to give lower F1 score, while nearly always improving the segmentation accuracy. These results likely mean there is a tradeoff between these two problems, despite coming from the same data. Overall, the model with an adapted classification branch, ResNet18 with 3 layers seems to give the best balanced results: 0.4245 mean AP with 93.00% COVID-19 sensitivity and 96.76% F1 score.

## 5 Conclusions

In this paper we have presented a fast and accurate single-shot architecture that both predicts the class of the image, COVID-19 vs Common Pneumonia vs Control, and segments lesions in the chest CT scans. Conceptual novelties include the addition of a separate classification branch within the RoI layer that outputs only encoded bounding boxes and their confidence scores that are used to train the image classification module. To the best of our knowledge, this is the first solution in the COVID-19 deep learning community that does a single-shot lesion instance segmentation and chest CT scan prediction.

We achieved the best results by training both branches from scratch: ResNet18 with the last truncated block achieves 0.4245 average precision for lesion segmentation, 93.00% COVID-19 sensitivity and 96.76% F1 score. Given the modest model of size of 8.27M parameters, of which only 1.9M are trainable during the classification stage, this model achieves the accuracy of a much larger classifier in [TS20b] with a ResNet50+FPN backbone and the segmentation precision of Mask R-CNN with ResNet50+FPN.

So far we have achieved strong results by only exploring the localization (bounding box) and the strength (confidence score) of the features. One of the key differences between COVID-19 and CP is the *configuration* of the lesions and features: diffuse distribution, attenuation, crazy-paving patterns, etc, that bounding boxes can’t capture. In our future research we intend to explore these configurations for COVID-19 prediction by considering the objects’ masks that capture the objects’ shapes more accurately than bounding boxes. The full source code, model interfaces and pretrained weights are available on https://github.com/AlexTS1980/COVID-Single-Shot-Model.

## Data Availability

http://ncov-ai.big.ac.cn/download

https://github.com/AlexTS1980/COVID-Single-Shot-Model

